# Automated IntraVascular UltraSound Image Processing and Quantification of Coronary Artery Anomalies: The AIVUS-CAA software

**DOI:** 10.1101/2025.02.18.25322450

**Authors:** Anselm W. Stark, Pooya Mohammadi Kazaj, Sebastian Balzer, Marc Ilic, Manuel Bergamin, Ryota Kakizaki, Andreas Giannopoulos, Andreas Haeberlin, Lorenz Räber, Isaac Shiri, Christoph Gräni

## Abstract

**Background:** Coronary artery anomalies (CAA) with an intramural course are associated with elevated risks of ischemia and sudden cardiac death under stress. Intravascular ultrasound (IVUS) is essential for assessing coronary vessel dynamics in these patients. However, the rarity of such anomalies, along with unique geometric changes in the intramural course and ostium, complicates image analysis, leading to inconsistencies and time-consuming evaluations. Our developed executable, zero/low-code software addresses these limitations by providing automated lumen segmentation and cardiac phase identification in IVUS images acquired during rest and stress protocols.

**Methods:** The software includes: (1) Automated segmentation of lumen contours trained and validated on 6713 frames (developed by using human in the loop active learning process) and tested on 914 frames, IVUS frames from fifteen patients (22 studies) with right CAA using a modified U2-Net deep learning (DL) model; (2) Extraction of systolic and diastolic frames via a dual-gating approach combining image- and contour-based methods; and (3) A graphical user interface enabling manual correction of the results. The gating module was validated using a custom flow-loop simulating patient-specific hemodynamics, while segmentation accuracy was assessed via intra-class correlation coefficient (ICC) analysis comparing AI-generated contours with those delineated by experienced readers.

**Results:** The DL model achieved a mean Dice score of 0.86 (SD: 0.07), sensitivity of 0.87 (SD: 0.11), and specificity of 0.98 (SD: 0.01) on the test set. ICC values for lumen area measurements were 0.92 (95%CI: 0.88-0.95) for rest and 0.98 (95%CI: 0.97-0.99) for stress conditions (all *p <* 0.001). The gating module demonstrated excellent reproducibility for identifying systolic and diastolic frames under both conditions (ICC = 1.00, *p <* 0.001).

**Conclusions:** AIVUS-CAA offers a reliable, automated tool for precise IVUS analysis at rest and during stress, enhancing the evaluation of geometrical changes of coronary vessels in CAA patients and enabling efficient clinical decision-making in a streamlined workflow.

**Graphical Abstract.**
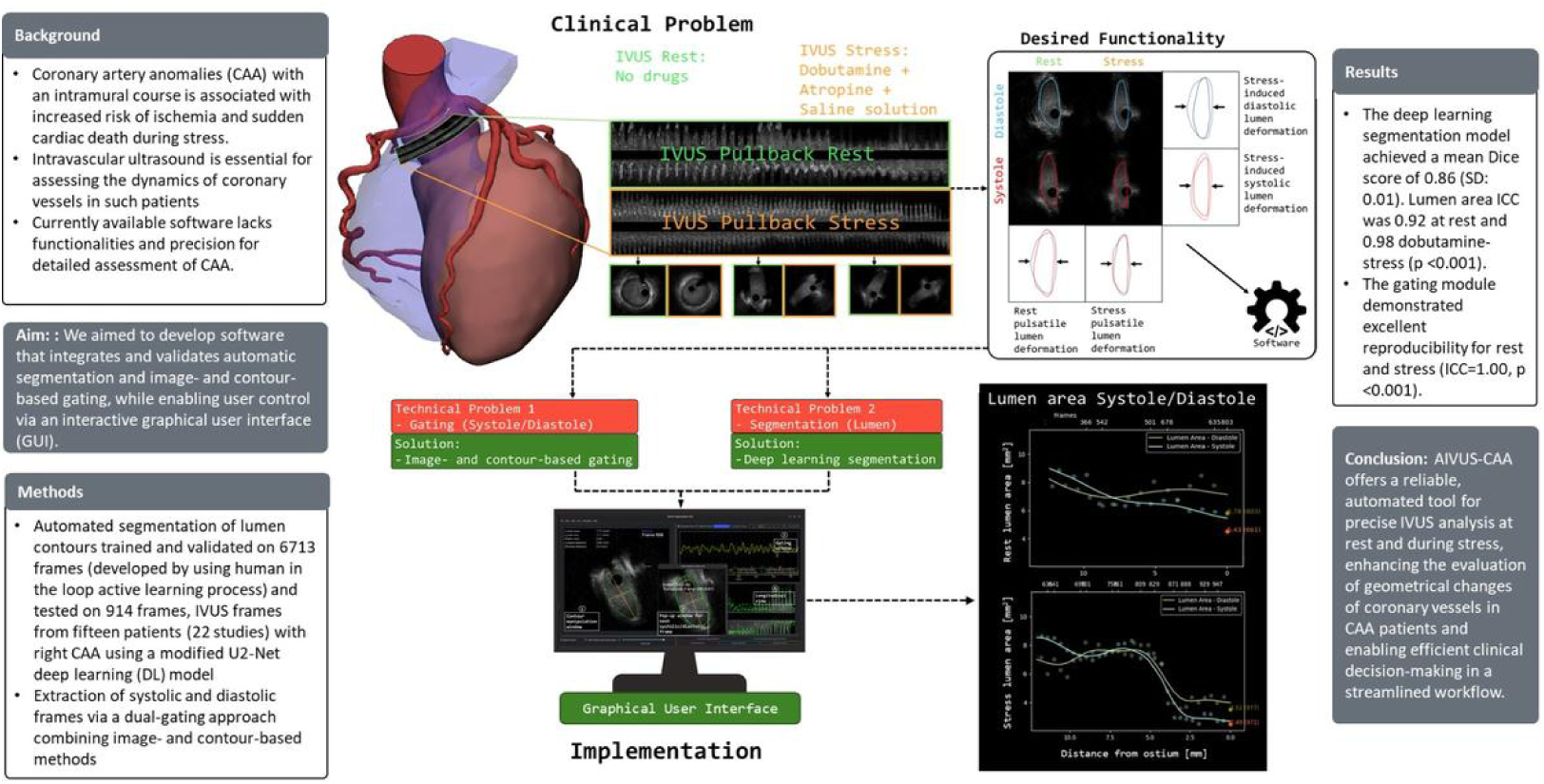

**Highlights:**

- Developed the first automated software (AIVUS-CAA) specifically designed to quantify dynamic geometric changes in coronary artery anomalies, addressing a critical gap in analyzing intramural coronary segments.
- Introduced a novel dual-gating algorithm combining image-based and contour-derived signals to identify systolic/diastolic phases, validated under both rest and simulated exercise conditions – a first for stress IVUS analysis in CAA.

## 1. Introduction

Coronary artery anomalies (CAA) are a rare congenital condition where an interarterial course (between the aorta and the pulmonary artery) and an intramural course (within the aortic wall) are of clinical interest, since these variants are associated with an elevated risk of ischemia and sudden cardiac death Eckart et al. (2004); Maron et al. (2009). The gold standard for hemodynamic assessment in patients with CAA is invasive coronary angiography combined with fractional flow reserve (FFR) and intravascular ultrasound (IVUS) during a dobutamine-atropine volume challenge, which mimics exercise conditions. This approach enables the capture of dynamic changes in coronary vessel geometric characteristic of CAA Bigler et al. (2021); Lee et al. (2016).

The vessel undergoes different geometric changes with changing states, first during every heartbeat either during rest (rest pulsatile lumen deformation) and then again during exercise condition (stress) (stress pulsatile lumen deformation). Additionally during stress, the intramural course of the vessel within the aortic wall is further deformed, during diastole (stress-induced diastolic lumen deformation) and more significantly during systole (stress-induced systolic lumen deformation). However, the underlying pathophysiology of CAA remains incompletely understood, partly due to the limitations of current IVUS analysis tools. Advanced IVUS analysis is essential for accurately interpreting the complex geometrical changes observed in these patients.

Comprehensive analysis of geometry and its change (rest/stress pulsatile lumen deformation and stress-induced diastolic/systolic lumen deformation) from IVUS requires software capable of: (1) automatically segmenting images acquired during pullbacks to handle the substantial volume of data and (2) identifying systolic and diastolic frames (i.e. gating) under both rest- Formato et al. (2023) and stress conditions, particularly within the intramural segment where the vessel geometry is mostly oval to slit-like and not round. Since most advanced IVUS systems are not synchronized with electrocardiograms (ECG), image-based gating algorithms were developed Talou et al. (2015); Torbati et al. (2019); Bajaj et al. (2021). However, these algorithms have been validated exclusively under rest conditions and in coronary artery disease not CAA. Stress protocols with increasing of heart rate and blood pressure, as applied in CAA patients, introduce additional complexity, especially for segmentation, due to the pronounced vessel motion during the cardiac cycle, which results in blurring and image artifacts. Moreover, it remains uncertain whether the geometrical changes observed in the intramural course allow the integration of contour-derived gating methods into a dual-gating framework (combining image- and contour-based approaches). Beyond dynamic analysis, precise anatomical measurements—such as lumen area, circumference, major and minor axis lengths, elliptic ratio, and the thickness of the aortic and pulmonary walls—are critical for understanding the mechanisms underlying complex vessel dynamics.

Currently, no commercially available tools offer the comprehensive functionality required for IVUS image analysis in CAA patients. To address this gap, we developed an executable, zero/low-code software that integrates automated segmentation and image-based gating, validated across both resting and stress conditions. Our solution also incorporates an interactive graphical user interface (GUI) to enable user-driven refinement of results.

## 2. Materials and Methods

### 2.1. Logic of Addressing Problems

To address the clinical challenge of quantifying rest/stress pulsatile lumen deformation and stress-induced diastolic/systolic lumen deformation in coronary vessels (Figure 1), we structured our approach around two core technical components: (1) segmentation and (2) gating. These processes are fully automated, enabling the system to identify systole and diastole phases and perform vessel segmentation without manual intervention. To provide users with additional control and refinement capabilities, we implemented a graphical user interface (GUI) (Figure 2) that allows for adjustments to the automated results. The methods section is organized as follows:

- **Data acquisition:** Description of the datasets and protocols used for training and testing.
- **Segmentation module:** Details of the automated lumen segmentation process.
- **Gating module:** Explanation of the dual gating approach to identify the systolic and diastolic frames.
- **Validation of the modules:** Methodology to evaluate the performance of the segmentation and gating modules.
- **Workbench module (GUI):** Overview of the user interface and its functionalities for manual corrections.
- **Software architecture specifications:** Summary of the system’s underlying technical architecture and implementation details.

**Figure 1:**
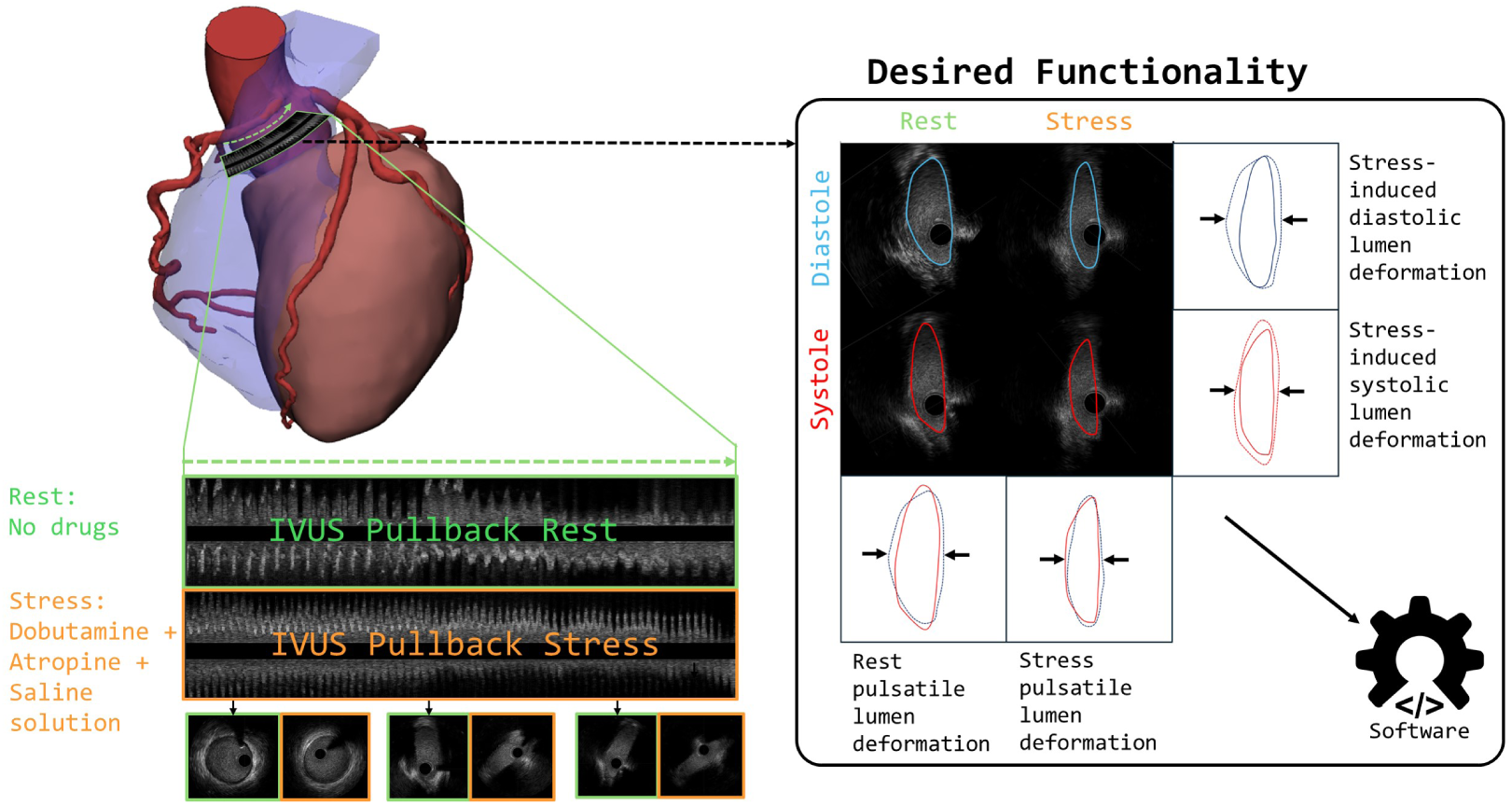
Clinical problem and desired functionality: On the left hand side, an acquired IVUS pullback through the intramural course of a CAA. This procedure is performed at two states: rest and simulated exercise-condition (stress). On the right side, the desired functionality to segment lumen area and identify systole and diastole to quantify lumen deformation from diastole to systole (rest pulsatile deformation and stress pulsatile deformation) and from rest to stress (stress-induced diastolic/systolic lumen deformation).

**Figure 2:**
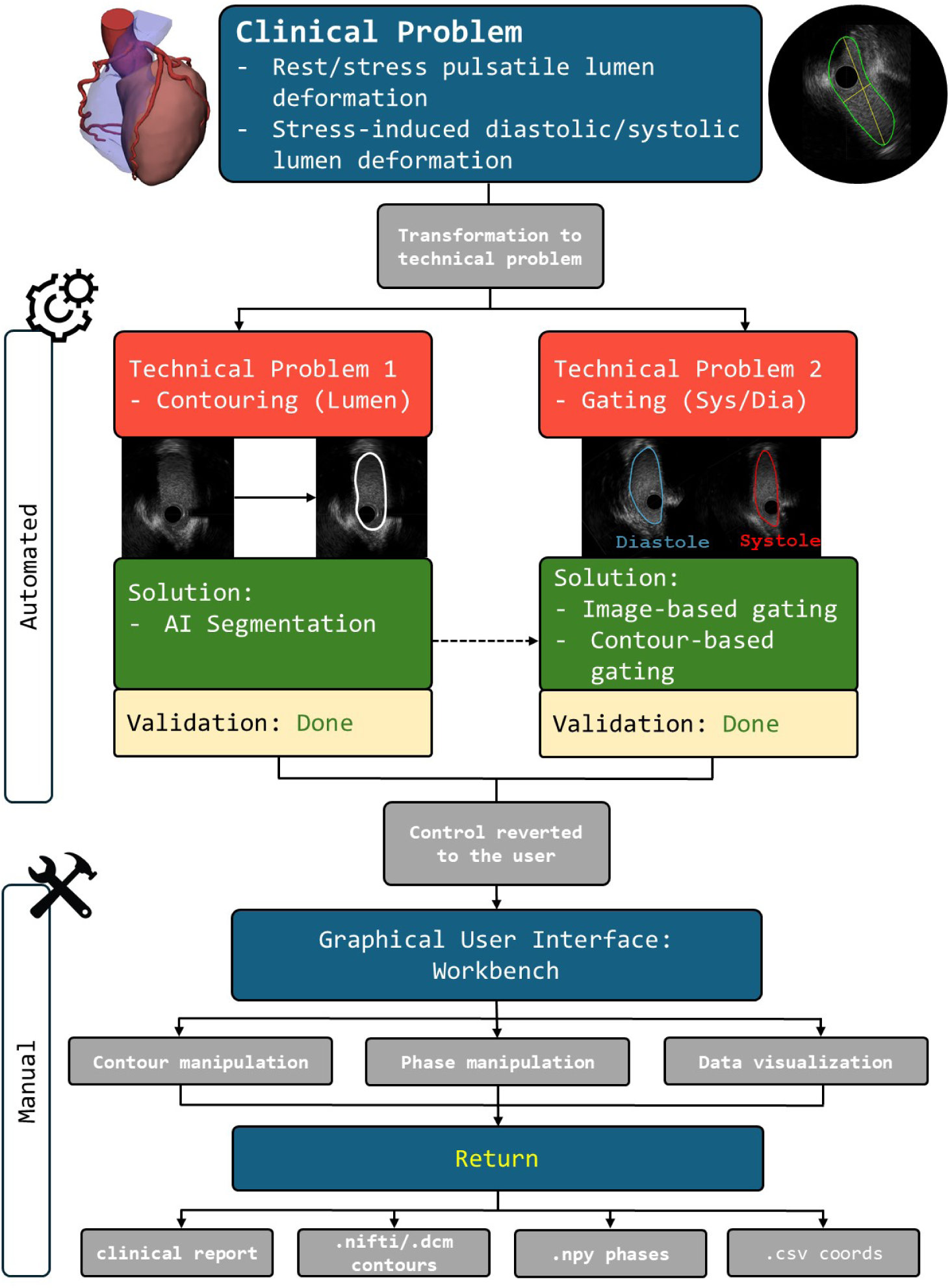
Flow chart of the CAA-specific IVUS processing pipeline. Segmented images are processed by the gating algorithm to identify frames of interest (i.e., systole and diastole). Control is then reverted to the user, enabling manual correction of the results. The software is designed to clearly distinguish between modules that allow user input and those that function automatically.

### 2.2. Data Acquisition

All IVUS DICOMs used for training the deep learning (DL) model and developing the gating module were sourced from our “Registry for Invasive and Non-invasive Anatomical Assessment and Outcome of Coronary Artery Anomalies (NARCO).” Ethical approval for the study was obtained (KEK 2020-00841), and all participants provided written, informed consent. The study is registered on ClinicalTrials.gov (ID: NCT04475289) Bigler et al. (2024).

For this software development project, we prospectively enrolled patients diagnosed with CAA featuring an interarterial and/or intramural course who presented at our specialized coronary artery anomaly clinic between June 2020 and October 2024. Inclusion criteria for the study were as follows: (1) the presence of any CAA with an interarterial and/or intramural course of the anomalous vessel, (2) age *≥* 18 years, and (3) provision of written informed consent. All patients underwent invasive coronary angiography and IVUS imaging during both rest and stress conditions.

Simulated exercise conditions were induced using a dobutamine-atropine-volume challenge protocol. Dobutamine was administered intravenously at an initial rate of 20 *µ*g/kg/min for 2 minutes, followed by 40 *µ*g/kg/min for 4 minutes. After 6 minutes, 0.5–1 mg of atropine was administered to achieve maximal heart rate (at least 85% of the predicted maximal heart rate or higher). To counteract the blood pressure reduction caused by dobutamine, 3 liters of saline solution were infused during the protocol.

IVUS imaging was performed using a standard protocol with automated pullback at a rate of 1 mm/s, employing a 40 MHz rotational transducer (Boston Scientific) capturing images at 30 frames per second, resulting in an interframe distance of 0.033 mm. The intraframe pixel spacing was 0.018 mm, providing a spatial resolution of 0.018 mm × 0.018 mm per pixel within each frame. Imaging was conducted during rest (*IV US_Rest_*) by positioning the IVUS probe distally to the intramural segment (i.e., in a region where the lumen was fully circular) and retracting the probe until only the aorta was visible. This process was repeated following the dobutamine-atropine-volume challenge (*IV US_Dobutamine_*).

A detailed description of the study protocol can be found in our methodology paper Bigler et al. (2024).

### 2.3. Segmentation Module

We implemented a human-in-the-loop active learning approach to tackle the time- and labor-intensive process of creating a dataset to train a segmentation model. Initially, we manually segmented 1,000 frames from three patients to develop a preliminary version of the network. This initial model was then applied to data from three additional patients with no pre-existing segmentations. From this dataset, we randomly selected 1,000 IVUS frames, which were subsequently reviewed by expert physicians specializing in IVUS imaging of CAA patients.

The physicians inspected the segmentations, selected the highest-quality ones, and modified them as necessary. They prioritized editing challenging cases where the network struggled to generate accurate segmentations. With each iteration, the focus was increasingly narrowed to more difficult cases (proximal section of coronary arteries). This iterative process was repeated until the model achieved satisfactory performance, where physicians no longer needed to modify segmentations for new cases. Using this approach, we generated a dataset comprising 7,627 frame-segmentation pairs from fifteen patients (22 studies). This dataset was then used to retrain and fine-tune the network.

For image segmentation, we utilized a modified U2-Net architecture with Residual U-blocks, a nested U-shaped design that enables multiscale feature extraction through mixed receptive fields. Deep supervision was employed to incorporate training loss across scales, improving local and global context extraction. The model was trained in 2D using the Adam optimizer with a learning rate of 0.001, L2-norm loss, a weight decay of 0.0001 and batch size of eight.

### 2.4. Gating Module

To identify systolic and diastolic frames, we employed a dual-gating approach that combines a modified image-based gating method with a contour-based gating approach. The contour-based approach is predicated on the assumption that the intramural course of the coronary artery undergoes significantly greater geometrical changes during rest pulsatile deformation than a normal coronary artery, making this approach potentially more valuable. Preprocessing of the images involved cropping each frame to a 400×400 pixel region centered on the IVUS catheter to eliminate frame labeling and extraneous details. To optimize performance speed, users can manually select specific regions of frames for processing.

#### 2.4.1. Image-Based Gating

For image-based gating, we employed a modified approach based on the method proposed by Maso Talou et al. and adapted for CAA by Formato et al. Talou et al. (2015); Formato et al. (2023). This approach utilizes pixel-wise correlation and blurriness metrics to detect phase transitions within the cardiac cycle. The method proceeds as follows.

First, the Pearson product-moment correlation coefficient (*ρ_t_*) is computed for each pair of consecutive IVUS frames (*I_t_* and *I_t_*_+1_) to quantify temporal consistency:

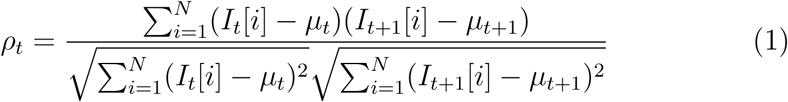

where *I_t_*[*i*] and *I_t_*_+1_[*i*] represent the pixel intensities at position *i* in frames *t* and *t* + 1, respectively, and *µ_t_* and *µ_t_*_+1_ are the mean pixel intensities of these frames. *N* denotes the total number of pixels in each frame.

A correlation coefficient *ρ_t_* close to *±*1 indicates high temporal consistency, typically corresponding to end-diastole or, to a lesser extent, end-systole, when the heart’s motion is minimal. In contrast, a *ρ_t_* close to 0 indicates substantial changes between the two frames.

For blurriness, we adapted the method of Maso Talou et al. by utilizing the Fast Fourier Transform (FFT) to analyze the frequency components of each frame Vysotska et al. (2024) instead of gradient magnitudes. The blurriness score (*B_t_*) for a given frame *I_t_* is calculated as follows: let *F* (*x, y*) represent the image in the spatial domain, and *ℱ* (*u, v*) be its 2D Fourier transform, where *u, v* are the frequency coordinates. The magnitude spectrum *M* (*u, v*) = *|ℱ* (*u, v*)*|* is computed, and the values are sorted in ascending order. The threshold is set at the 90% percentile of the sorted values, as determined through testing. The blurriness score is derived as the mean of the top 10% of the magnitudes (corresponding to the highest frequencies):

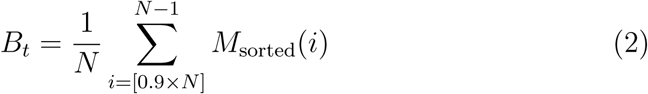

Higher *B_t_* values indicate sharper images with more high-frequency components, whereas lower *B_t_* values suggest increased blurriness, typically due to heart motion.

The correlation coefficient (*ρ_t_*) and blurriness score (*B_t_*) were normalized using z-scores. A Butterworth band-pass filter was applied to both signals to smooth the data and eliminate noise outside the physiological heart rate range. The filter was designed with a low cutoff frequency of 1.33 Hz and a high cutoff frequency of 6.0 Hz, corresponding to twice the expected heart rate range (80 bpm to 360 bpm) observed during our invasive procedures. This adjustment accounts for the fact that systolic and diastolic peaks are detected separately.

Local maxima were identified for both metrics, and the variability of the maxima intervals was assessed by calculating the variance of the intervals between consecutive maxima. A combined signal (*s*_image_(*t*)) was generated using a weighted combination of the normalized signals, where the weights are inversely proportional to the variability of each signal:

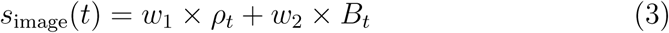

where:

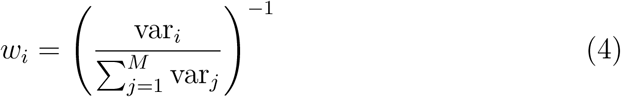

In this formula, *M* represents the total number of signals being combined, and var*_j_* is the variance of each individual signal. This approach assumes that heart rate variability remains relatively constant during the image acquisition period.

#### 2.4.2. Contour-Based Gating

Contour-based gating leverages geometric dynamics derived from segmentation data. For the clinical report, several key metrics are calculated from the segmented images: lumen area, lumen circumference, elliptic ratio, and both the longest and shortest distances. Additionally, a vector is computed from the center of the image (i.e., the catheter) to the centroid of the contour, which enables tracking of vessel movement over time by monitoring the vector’s angle and length. Preliminary analysis revealed that the shortest distance (SD), vector length (VL), and vector angle (VA) were the most sensitive to temporal changes (Figure A.7).

These three signals (SD, VL, and VA) were then normalized using z-scores, and a Butterworth bandpass filter was applied to remove noise and smooth the signals. Local extrema were identified, and their variability was assessed to determine the appropriate weights for the combined signal. The combined contour signal is given by:

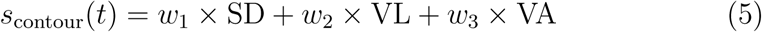

where:

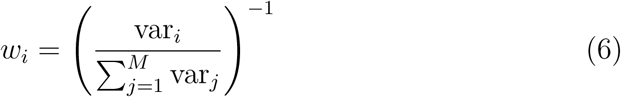

Here, var*_i_* represents the variance of the individual signal, and *M* is the total number of signals combined.

For automatic gating using the dual-gating approach, the diastolic and systolic phases are identified by analyzing both the image-derived signal (*s_image_*) and the contour-derived signal (*s_contour_*). Specifically, the phases are defined where the peaks of these two signals overlap most closely. A phase is set only when the peaks of both signals align sufficiently. Additionally, an optional third signal can be incorporated by specifying the initial diastolic and systolic frames (from the proximal region). These frames are correlated with other frames within a defined range (i.e., *±* buffer frames) based on the estimated heart rate, which is derived from the distances between the maxima (or extrema) of both *s_contour_* and *s_image_*. The frame with the highest Pearson product-moment correlation coefficient is identified as the reference, and this process is iteratively applied to each newly identified “highest-correlation” frame.

### 2.5. Validation of segmentation and gating module

#### 2.5.1. Validation of Segmentation Module

The model was internally validated using standard evaluation metrics in validation and hold-out test set (patient-wise splitting), including Dice score, sensitivity, and specificity, to assess the performance of the automatic segmentation algorithm.

To evaluate the segmentation accuracy, three random patients were selected for analysis. For each patient, 16 frames from the distal vessel segment and 16 frames from the intramural vessel segment were chosen under both rest and stress conditions, totaling 96 frames for rest and 96 frames for stress conditions (192 frames in total). The segmentation was first performed by the AI model and then manually by an experienced reader.

The following parameters were used for comparison: lumen area and SD. The AI segmentation results were compared to the manual segmentation for each frame, using Dice score, sensitivity, and specificity. The analysis was conducted first for all frames combined (independent of rest or stress conditions) and then separately for rest and stress conditions.

An Intraclass Correlation Coefficient (ICC) was also computed for each set of parameters (lumen area and SD). An ICC value greater than 0.90 was considered indicative of excellent performance, and a value between 0.75 and 0.90 was considered indicative of good performance.

#### 2.5.2. Validation of Gating Module

The gating module was validated using a custom-built flow-loop, as previously described Illi et al. (2025). Patient-specific hemodynamic parameters, corresponding to the time of IVUS acquisition during both rest and stress, were recreated individually using the flow-loop. An IVUS pullback was then performed in the same manner as in the clinical study.

For registration, an indentation was created in the distal vessel by applying external pressure A.8. This indentation, which caused an elliptical deformation visible in the IVUS images, corresponded to a pressure drop in the distal pressure curve measured by FFR. These two regions of interest, the elliptical deformation in the IVUS images and the drop in the pressure curve inside the vessel (distal pressure), were used to register the IVUS images and the pressure curves (Figure 3).

**Figure 3:**
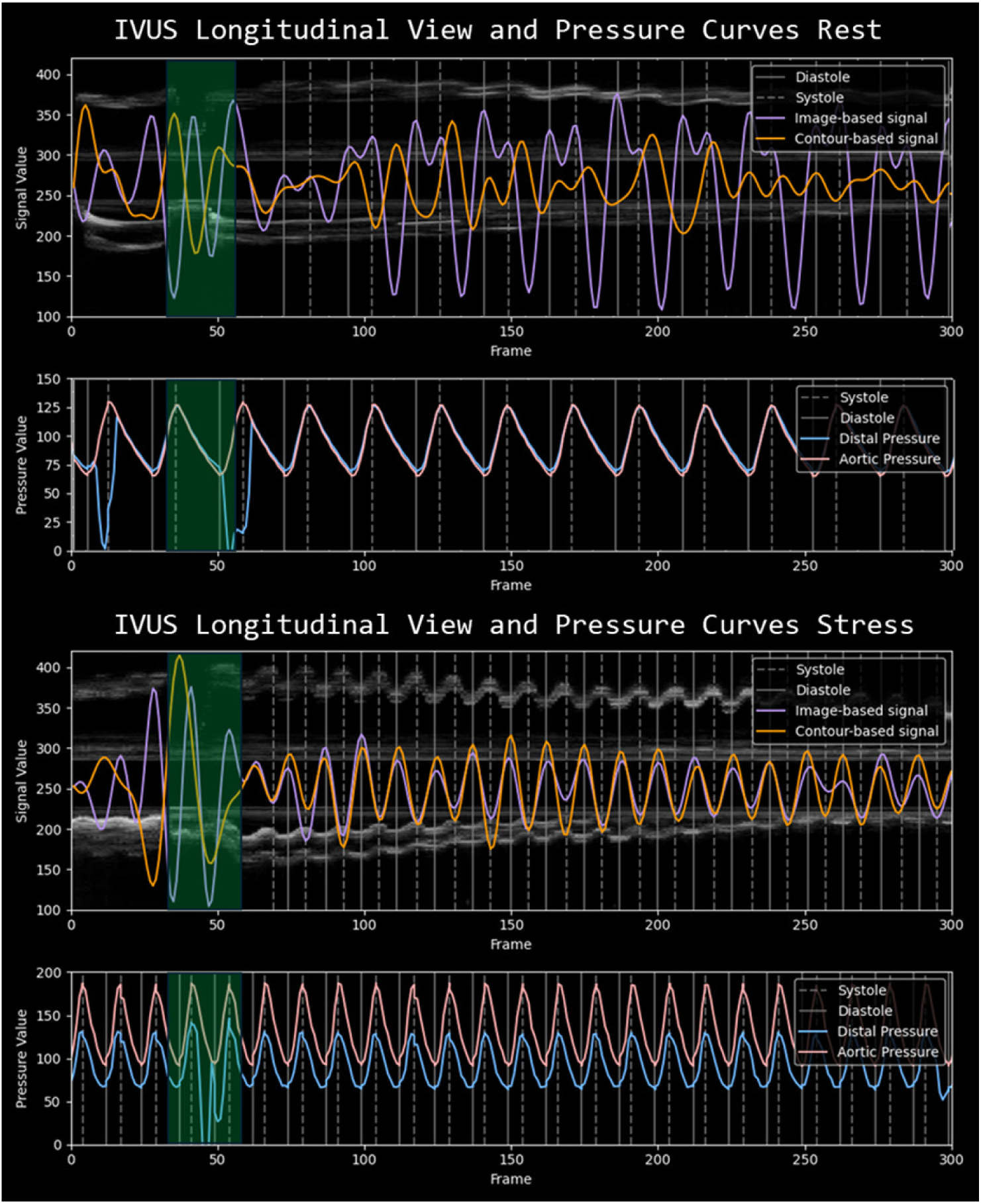
The two images at the top represent rest conditions, while the two images at the bottom correspond to stress conditions. In both cases, the upper images show the longitudinal view of the IVUS images with the extracted signals s_image_ and s_contour_, as well as the IVUS-derived diastolic and systolic phases. The lower images display the corresponding pressure curves with marked diastolic and systolic points. The images are registered using an indentation created in the vessel phantom, which is also visible in the pressure curve (highlighted by a green overlay).

Systolic and diastolic frames were identified within the IVUS pullback using the gating module (with manual adjustment). A peak detection algorithm was applied to the pressure curves to locate the corresponding systolic and diastolic points.

To evaluate the accuracy of the gating module, an ICC analysis was performed to compare the number of frames between systolic and diastolic points in the pressure curves and the corresponding IVUS frames. Additionally, ICC analysis was conducted to assess agreement between tool-assisted gating and visually determined gating based on raw IVUS images. Finally, inter-reader ICC was calculated for three randomly selected patient cases to assess consistency between two independent experienced readers.

All statistical analyses were performed using Python (version 3.10.12) and R (version 4.2.3).

### 2.6. Workbench Module (GUI)

The workbench module of the software provides an interactive platform through a graphical user interface (GUI) that enables users to correct and refine the results of automatic segmentation and gating (Figure 2).

The GUI is divided into two sections: the left side contains the functionality of the segmentation module, and the right side contains the functionality of the gating module (Figure 4).

**Figure 4:**
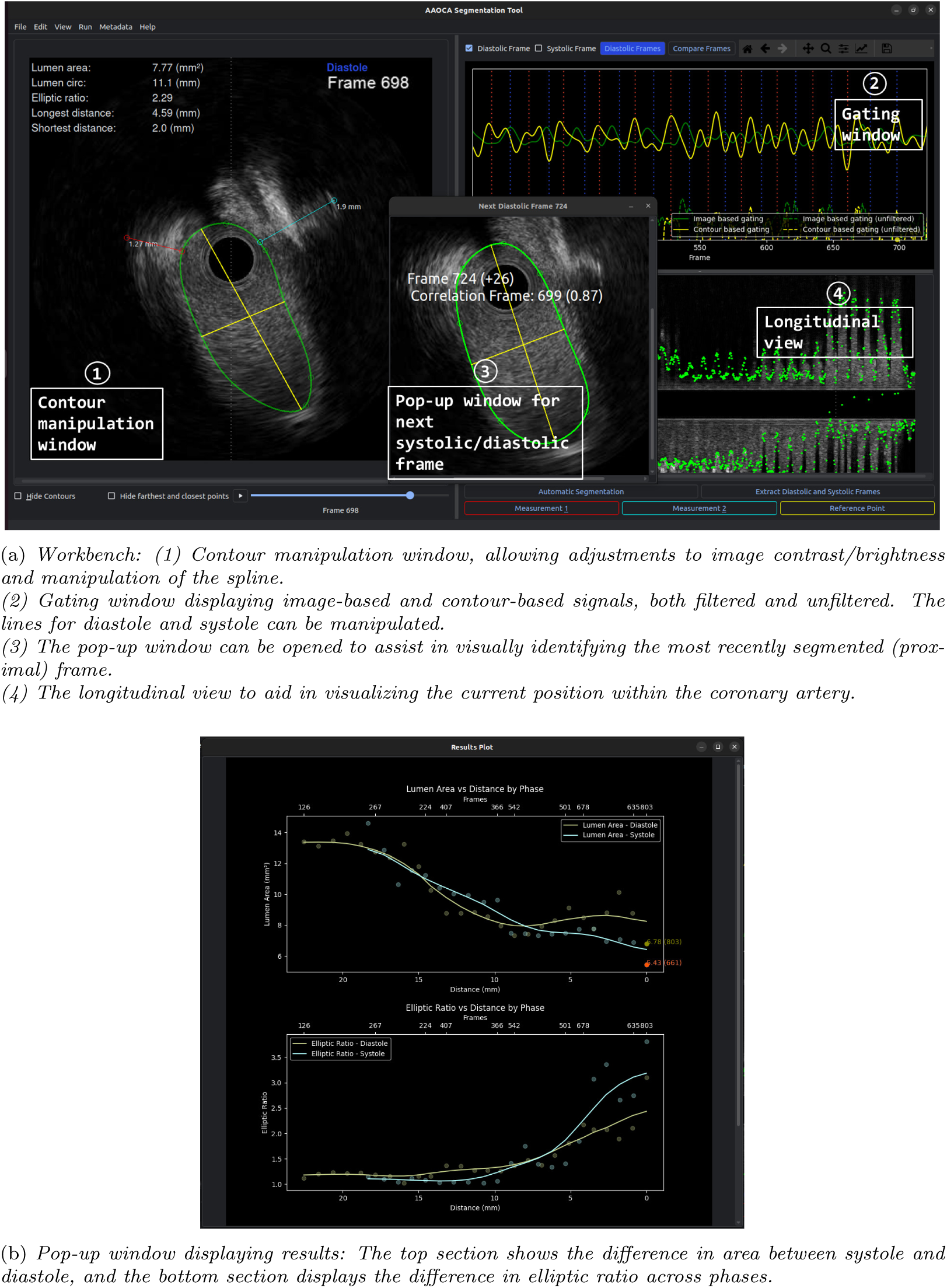
Graphical user interface windows. a) Main window (with small pop-up window) b) Pop-up window with results.

In the contour manipulation window (Figure 4, number 1), the image brightness, contrast, and color can be adjusted. The mask is transformed into a spline object, which can be manually manipulated by dragging points or adding new ones. This feature allows users to fine-tune the AI-generated segmentation. Additionally, measurement lines can be drawn to assess the distance between the CAA lumen and the aorta, as well as between the CAA and the pulmonary artery (Figure 4, number 1; red line for aortic wall thickness and blue line for pulmonary artery).

The gating window (Figure 4, number 2) displays the previously calculated *s_image_* and *s_contour_* signals, both filtered and unfiltered, along with the automatically labeled systolic/diastolic frames. These lines can be freely moved, deleted, or added. For additional assistance, a pop-up window can be opened to display the next proximal frame and the suggested next distal frame with the highest Pearson product-moment correlation coefficient (Figure 4, number 3).

A detailed description of the GUI functionalities is provided in a tutorial video on our GitHub page: AI in Cardiovascular Medicine - AIVUS-CAA Repository.

### 2.7. Software Architecture and Specifications

The software was developed in Python (version 3.10.12) and tested on a Linux operating system (Ubuntu 22.04.5). The GUI is built using PyQt5. The architecture is modular, allowing for easy integration of additional modules by other developers.

The project follows a well-organized directory structure that separates the code into distinct components to ensure clarity and maintainability (Supplemental Figure A.12). A full list of dependencies is provided in the ‘pyproject.toml’ file, which is automatically handled by Poetry to simplify installation and management.

To facilitate the integration of new features, the software structure supports a clear separation of functionalities. For example, the gating and segmentation modules are independent, enabling easy modification or extension of these features without affecting other parts of the software. The modular design allows developers to easily add custom modules for tasks such as advanced image segmentation or custom gating methods.

The software accepts standard DICOM images from all major vendors and .nifti files as input. For output, the software generates a clinical report in .txt format, provides geometrical measurements in .json format, exports the contours in .nifti format or .dcm format for model training, and stores the gated frames as .npy files for efficient post-processing of DICOM pixel data. These file formats are widely used in medical imaging and machine learning workflows, but can be easily extended to meet user-specific needs.

Users with compatible hardware can benefit from improved computational efficiency when running the software on Linux with GPU support.

The full source code is available at the following GitHub repository: AI in Cardiovascular Medicine - AIVUS-CAA Repository.

## 3. Results

### 3.1. Validation of Segmentation

The DL model achieved a mean Dice score of 0.86 (SD: 0.07), sensitivity of 0.87 (SD: 0.11), and specificity of 0.98 (SD: 0.01) on the test set. The ICC analysis conducted to validate the automatic segmentation tool against an experienced reader showed excellent agreement. During resting conditions, the ICC value for the absolute agreement of a single rater for the area was 0.90 (95% CI: 0.86-0.93) and the absolute ICC of the average raters was 0.95 (95% CI: 0.92-0.97). For the shortest distance, the absolute ICC of the individual rater was 0.95 (95% CI: 0.92-0.97) and the absolute ICC of the average raters was 0.97 (95% CI: 0.96-0.98).

Under stress conditions, the absolute ICC value of the single rater for the area was 0.98 (95% CI: 0.96-0.98), and the absolute ICC value of the average raters was 0.99 (95% CI: 0.98-0.99). For the shortest distance, the absolute ICC of the individual rater was 0.99 (95% CI 0.98-0.99) and the absolute ICC of the average rater was 0.99 (95% CI 0.99-1.00) (Figure 5a).

**Figure 5:**
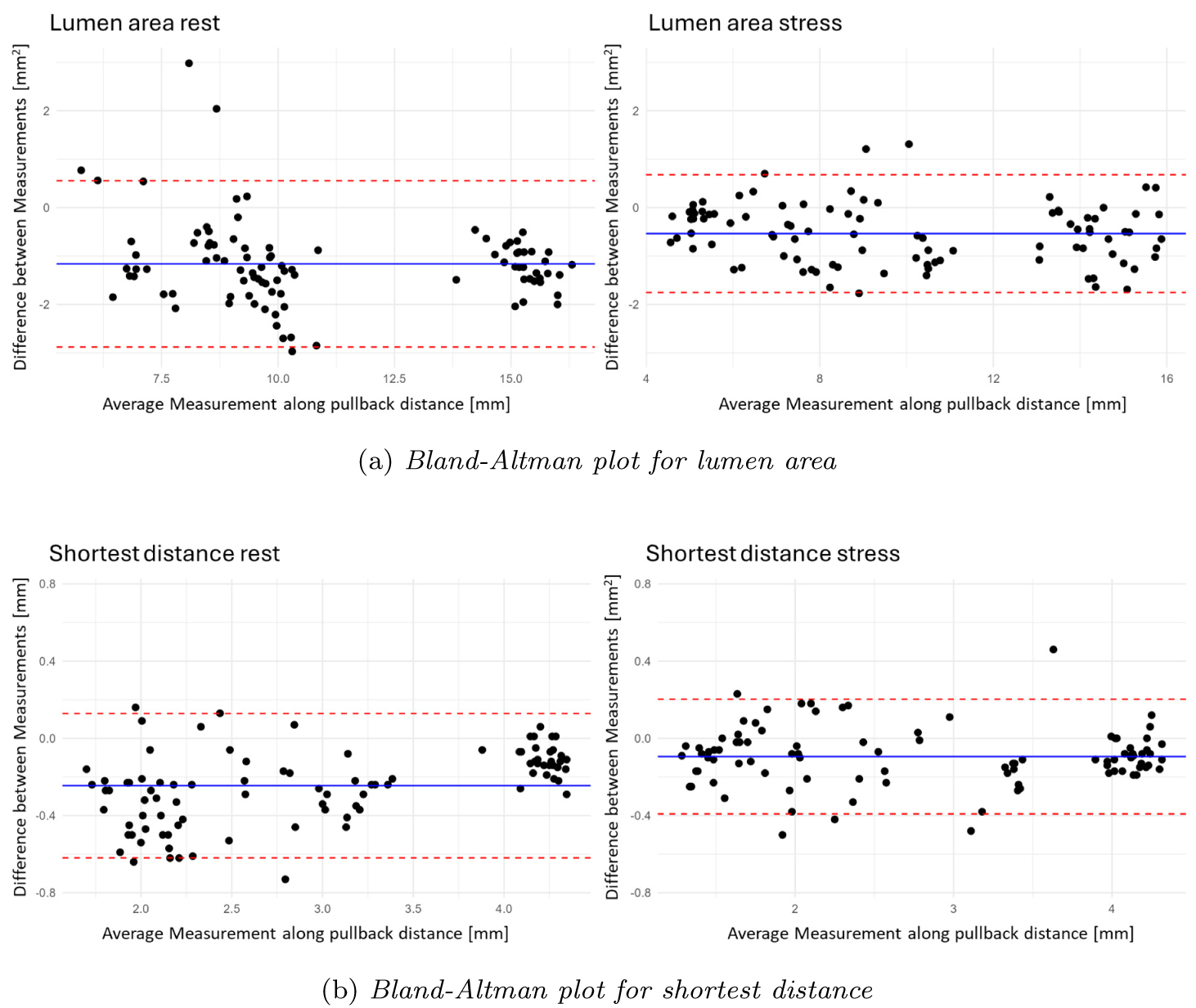
(a) Bland-Altman plots comparing lumen area of automatically segmented IVUS images and user-contoured frames. (b) Bland-Altman plots comparing shortest distance of automatically segmented IVUS images and user-contoured frames.

The inter-reader ICC values for the area were 0.94 (95%CI: 0.87-0.97) for rest and 0.91 for stress (95%CI: 0.85-0.98). For shortest distance, the ICC values were 0.97 (95%CI: 0.85-0.98) for rest and 0.98 (95%CI: 0.97-0.99) for stress (Supplemental Figure A.11).

### 3.2. Validation of Gating

The ICC analysis conducted to validate the gating module against a custom flow-loop demonstrated excellent reproducibility in identifying systolic and diastolic frames under both rest and stress conditions. For the rest phase, both systolic and diastolic frames yielded ICC values of 1.00 (95% CI: 1.00-1.00) for both tool-assisted and manual (”visual”) methods (*p <* 0.001). During the stress phase, the results were similarly robust, with ICC values remaining at 1.00 (95% CI: 1.00-1.00) for both systolic and diastolic frames under all conditions, for both tool-assisted and visual methods (*p <* 0.001). Bland-Altman plots showed that the tool-assisted gating demonstrated a smaller 95% CI and less systematic bias compared to the ground truth of the pressure measurements, particularly for resting conditions (Figure 6). Notably, manual correction was required for most phases, especially when applied to real-life cases as opposed to the idealized testing environment.

**Figure 6:**
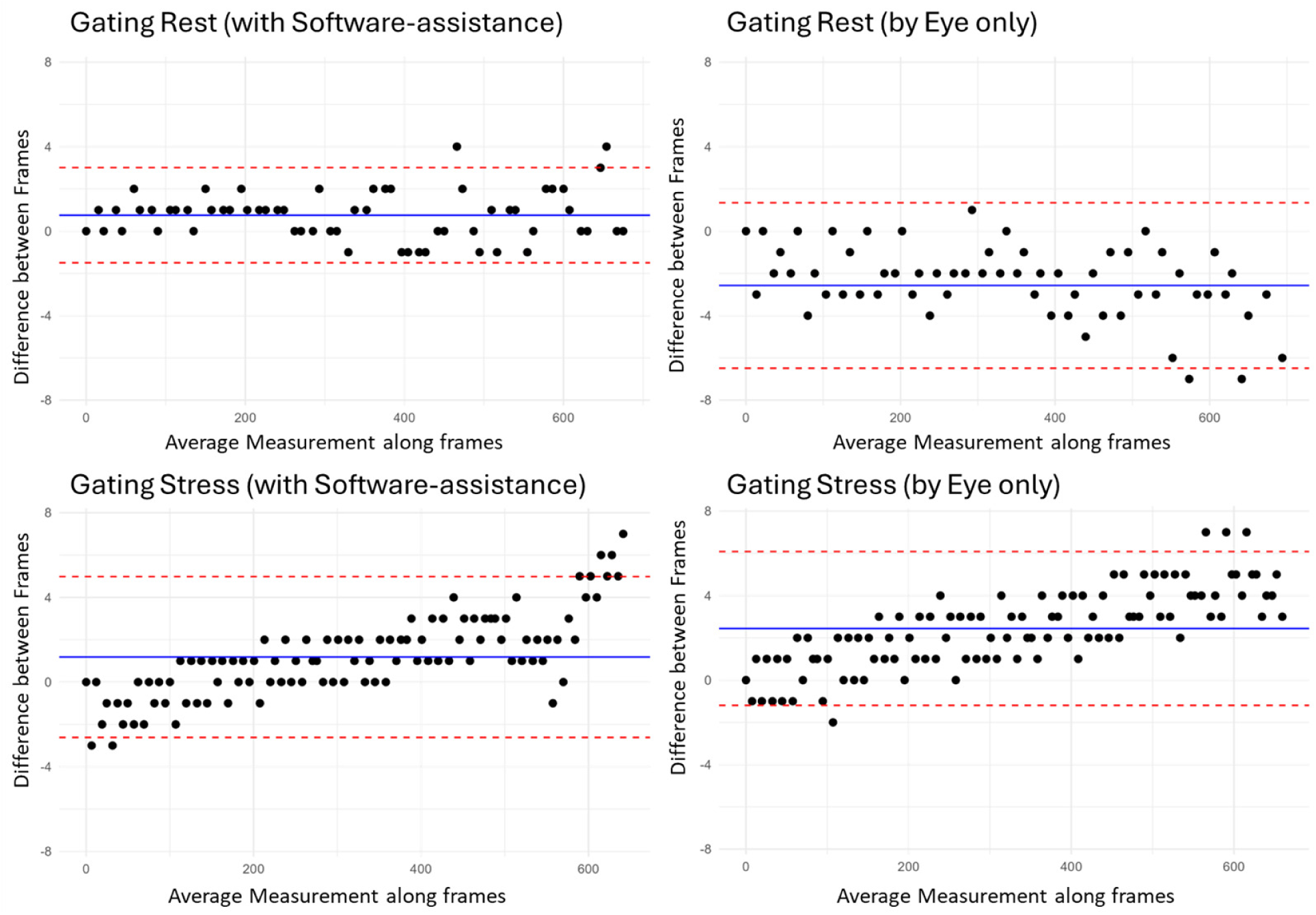
Comparison of gating using the software versus manual (”visual”) analysis. The top plot shows the difference during rest, and the bottom plot shows the difference during stress.

Intra-reader analysis showed that both the gating tool and manual analysis achieved excellent performance, with ICC values of 1.00 (95% CI: 1.00- 1.00). However, manual analysis exhibited a larger 95% CI (Supplemental Figure A.9).

Inter-reader analysis for gating can be found in Supplemental Figure A.10.

## 4. Discussion

This study introduces a novel, comprehensive software tool designed to enhance the analysis of coronary vessel dynamics in patients with CAA. The tool specifically quantifies rest/stress pulsatile lumen deformation and stress-induced diastolic/systolic lumen deformation. Our DL model for automatic lumen segmentation demonstrated excellent performance when compared to experienced readers. Additionally, our dual-gating approach showed excellent results when validated against data acquired from a custom flow-loop, exhibiting slightly less variation compared to visual assessment. However, the software-assisted gating analysis was found to be less time-consuming and more reproducible.

Several studies have proposed automatic segmentation methods for IVUS images Jeong et al. (2024); Kim et al. (2024); Nishi et al. (2020); Dong et al. (2023); Bajaj et al. (2021); Zhu et al. (2022). However, none have specifically focused on CAA. These models yielded similar Dice scores Dong et al. (2023); Kim et al. (2024); Jeong et al. (2024). Dong et al. used ICC values in their dataset of 11,070 images to compare different anatomical measurements, resulting in a strong agreement comparable to our results Dong et al. (2023). However, there is a clear need for DL models specifically trained on CAA data. The geometry of coronary vessels inside the aortic wall differs significantly from that of normal coronary arteries, both with and without coronary artery disease. The borders are also less clearly defined in the presence of calcification or healthy vessels, leading to more challenging segmentation tasks, as highlighted by our data. Notably, our DL model achieved ICC results comparable to those obtained between two expert readers while also significantly reducing analysis time.

The gating approach in our software was primarily based on the method proposed by Maso Talou et al. Talou et al. (2015), which was further adjusted by Formato et al. to analyze systolic and diastolic changes in CAA Formato et al. (2023). The original approach relied solely on an image-based gating method to extract the diastolic phase, which was extended by Formato et al. to also extract the systolic phase. Our unique contribution lies in the incorporation of contour-based gating methods, creating a dual-gating approach. Although our gating module demonstrated strong performance when validated against flow-loop data, manual correction improved reliability compared to fully automatic detection. The accuracy of the automatic approach varied depending on imaging quality, prompting us to focus primarily on manually corrected phases, which showed excellent agreement with flow-loop ground truth. While the tool did not significantly outperform visual phase assessment, it improved reproducibility, precision, and most importantly, reduced analysis time. Additionally, Bland-Altman plots suggested that manual gating introduced a higher bias, with a mean value farther from zero compared to tool-assisted gating. This improvement is likely due to the fact that the tool assists physicians in making data-driven decisions by plotting both image-derived and contour-derived information, thereby improving accuracy. However, there does not appear to be a one-size-fits-all approach, as the signal data is highly dependent on factors such as state (rest or stress), image quality, and other unknown variables. Additional DL approaches could potentially streamline automatic detection in the future Bajaj et al. (2021); Huang et al. (2023). Future versions of the software could also explore the integration of different image-based gating algorithms, which have shown potential in coronary artery disease Torbati et al. (2019). Notably, our ICC analysis of inter-reader reliability demonstrated high reproducibility, facilitating the acquisition of consistent and reliable data for further analysis of rest/stress pulsatile deformation and stress-induced diastolic/systolic lumen deformation.

Interestingly, Huang et al. demonstrated that a neural network-based approach achieved approximately 70% accuracy, surpassing human readers, underscoring the complexity of this task Huang et al. (2023). Our unique semi-automatic approach, which integrates additional data beyond what is visually perceptible, shows promise for addressing such challenges. Furthermore, with continued improvements to our DL model, the contour-based gating approach could be further optimized.

This tool could greatly assist clinicians in diagnosing CAA and quantifying dynamics (rest/stress pulsatile lumen deformation and stress-induced diastolic/systolic lumen deformation), especially in cases where traditional methods might struggle due to the atypical geometry of the vessels. The improved reproducibility and precision provided by the software may reduce inter-reader variability, thus potentially leading to more accurate and consistent clinical assessments.

## 5. Limitations

One limitation of our study is that the data was not ECG-gated, and validation against ECG-gated data was not performed. However, we incorporated a custom flow-loop phantom, allowing us to assess accuracy under controlled conditions. While this provides a strong foundation, additional validation using clinical ECG-gated IVUS datasets, particularly in patients with irregular heart rhythms, will be necessary for broader applicability.

Another limitation is that our software was primarily tested on right-sided anomalous aortic origin of coronary arteries with an intramural course. Although this represents a critical use case, further testing across a wider range of coronary anomalies and pathologies is needed to enhance generalizability. Furthermore, Python, as the underlying programming language, may introduce performance constraints when handling large datasets or real-time processing. However, our primary focus was ensuring segmentation accuracy and gating reliability.

Additionally, detecting systole and diastole alone may not fully capture rest/stress pulsatile lumen deformation and stress-induced diastolic/systolic lumen deformation, as external factors such as breathing motion can influence catheter positioning. Future enhancements should incorporate methodologies to account for these variables, improving real-world applicability.

Lastly, while a Windows version of the software has been developed, it does not currently support GPU acceleration, leading to lower performance. This limitation can be mitigated by using the Windows Subsystem for Linux (WSL) or virtual machines.

## 6. Conclusion

In conclusion, this study introduces a novel, modular, zero/low code, and executable software that significantly enhances the analysis of coronary artery anomalies, particularly in patients with CAA. Our DL-based segmentation and dual-gating approach have demonstrated excellent performance in both validating the lumen area and identifying systolic and diastolic frames with high precision. The software’s ability to improve reproducibility and reduce inter-reader variability positions it as a promising clinical tool for the diagnosis and assessment of complex vessel dynamics. Despite its current limitations, the tool provides a substantial step forward in the development of advanced imaging analysis for coronary artery anomalies.

## Data Availability

The raw datasets are protected and are not available due to data privacy laws. Access can be obtained upon IRB and Data Sharing Committee approvals from Bern, and the requesting institution, within a time frame of one year upon reasonable request to the authors

## Appendix A. Appendix Section

**Figure A.7:**
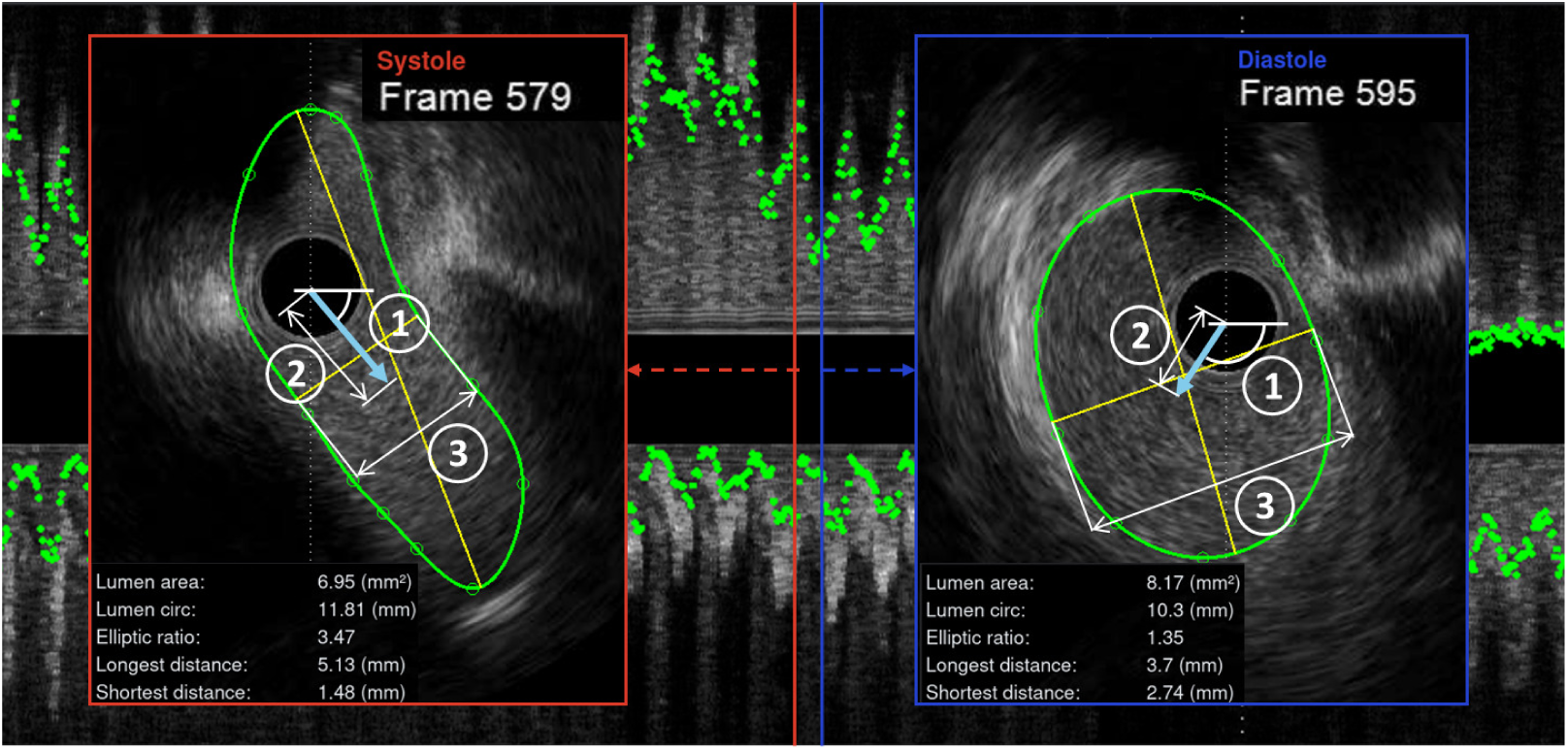
The different contour measurements used for the contour-based gating. Number 1 shows the angle of a vector pointing from the catheter to the centroid of the contour and number 2 represents the vectors length. Number 3 is the shortest distance. On the left side in the red box are the measurements for a systolic frame, on the right are the same measurements during diastole for a neighboring phase.

**Figure A.8:**
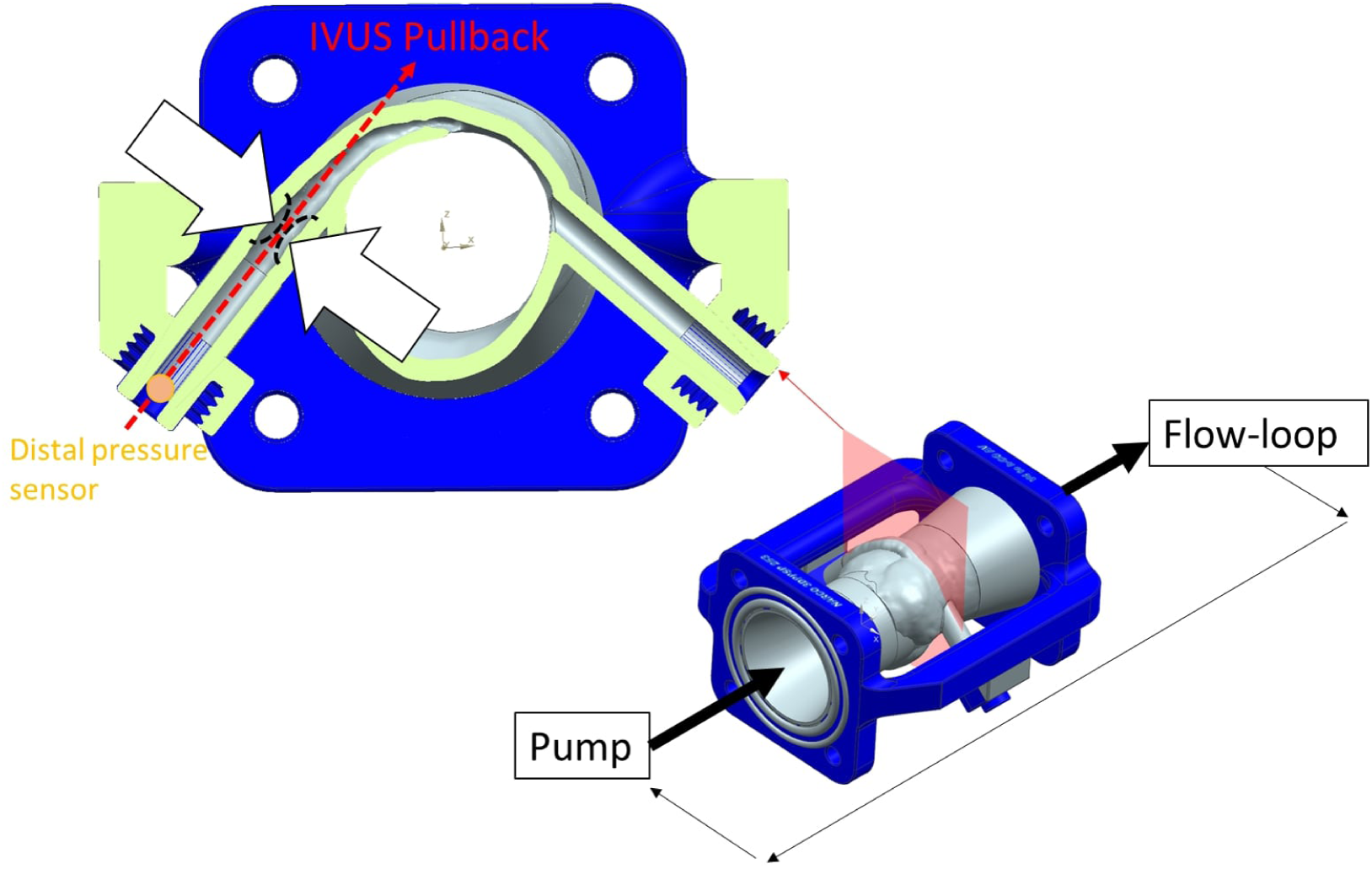
Adjusted from Illi et al. “Hemodynamic Relevance Evaluation of Coronary Artery Anomaly During Stress Using FFR/IVUS in an Artificial Twin”Illi et al. (2025): showing the transvertional cut through the vessel phantom and the position of the vessel indentation (white arrows) as well as the direction of the IVUS pullback (red dotted line) and the distal pressure sensor (orange point).

**Figure A.9:**
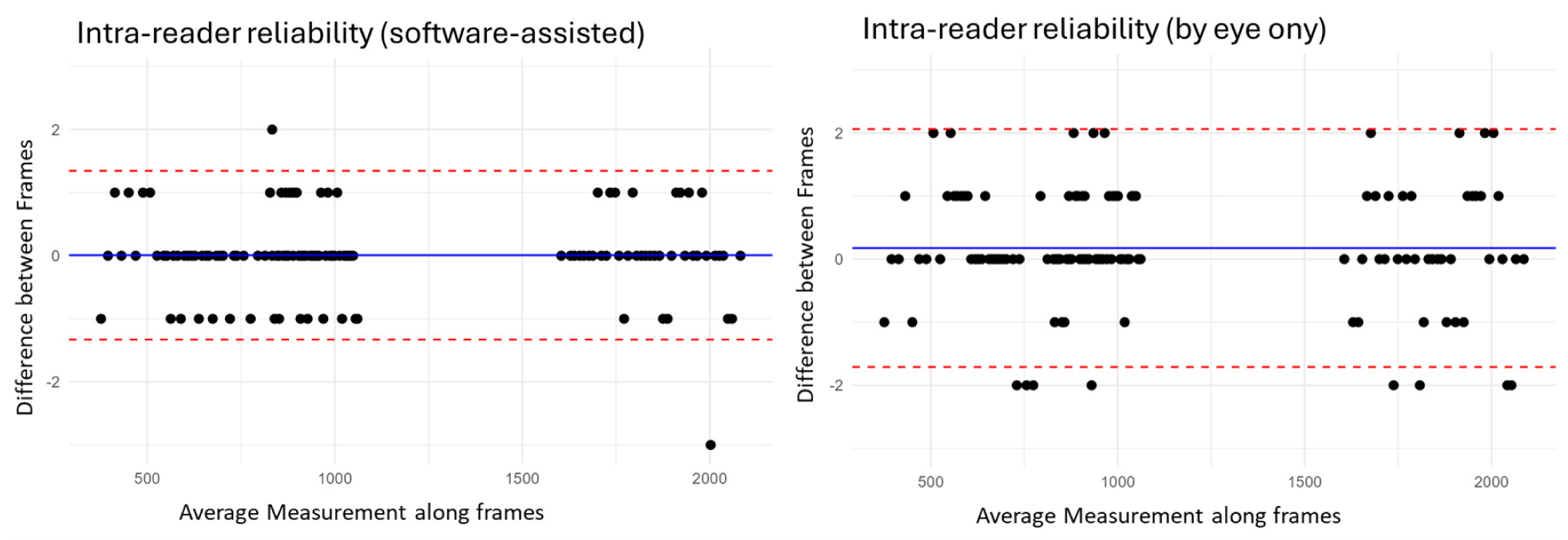
Comparison of software-assisted gating against solely visually. On top, the difference during rest and on the bottom during stress.

**Figure A.10:**
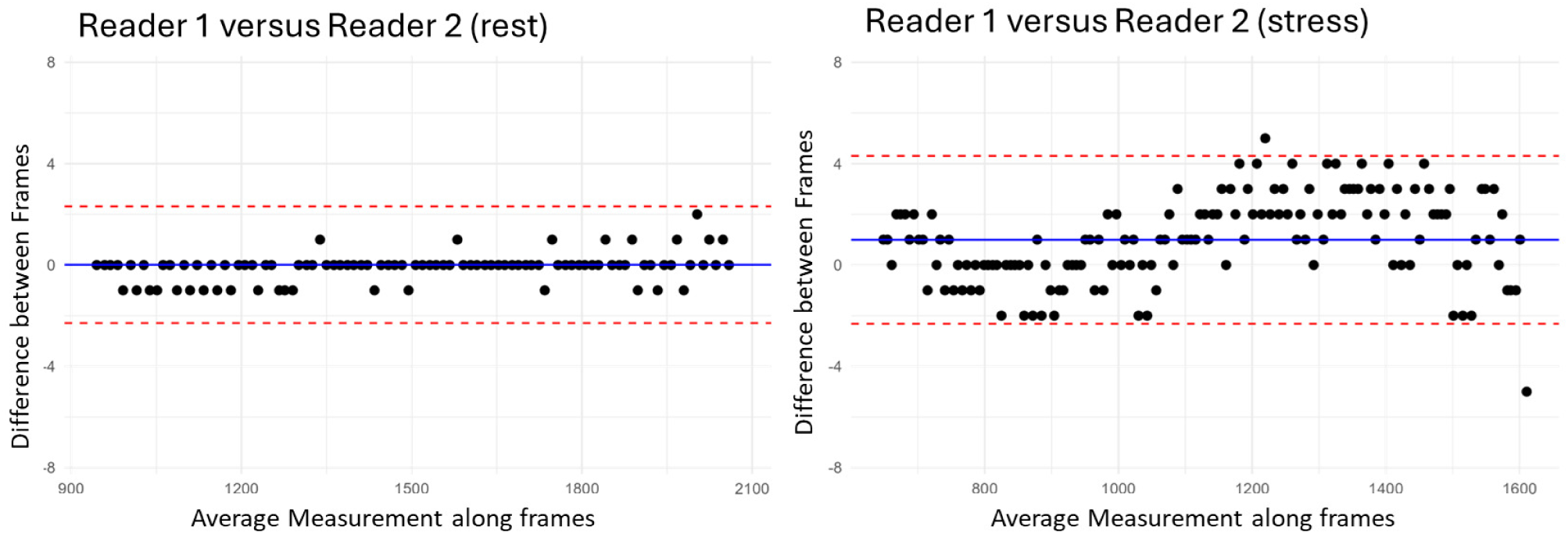
Bland-Altman plot for showing the difference between reader 1 and reader 2 for tool-assisted gating. The ICC was 1.00 (95%CI: 1.00-1.00, p < 0.001) for both rest and stress.

**Figure A.11:**
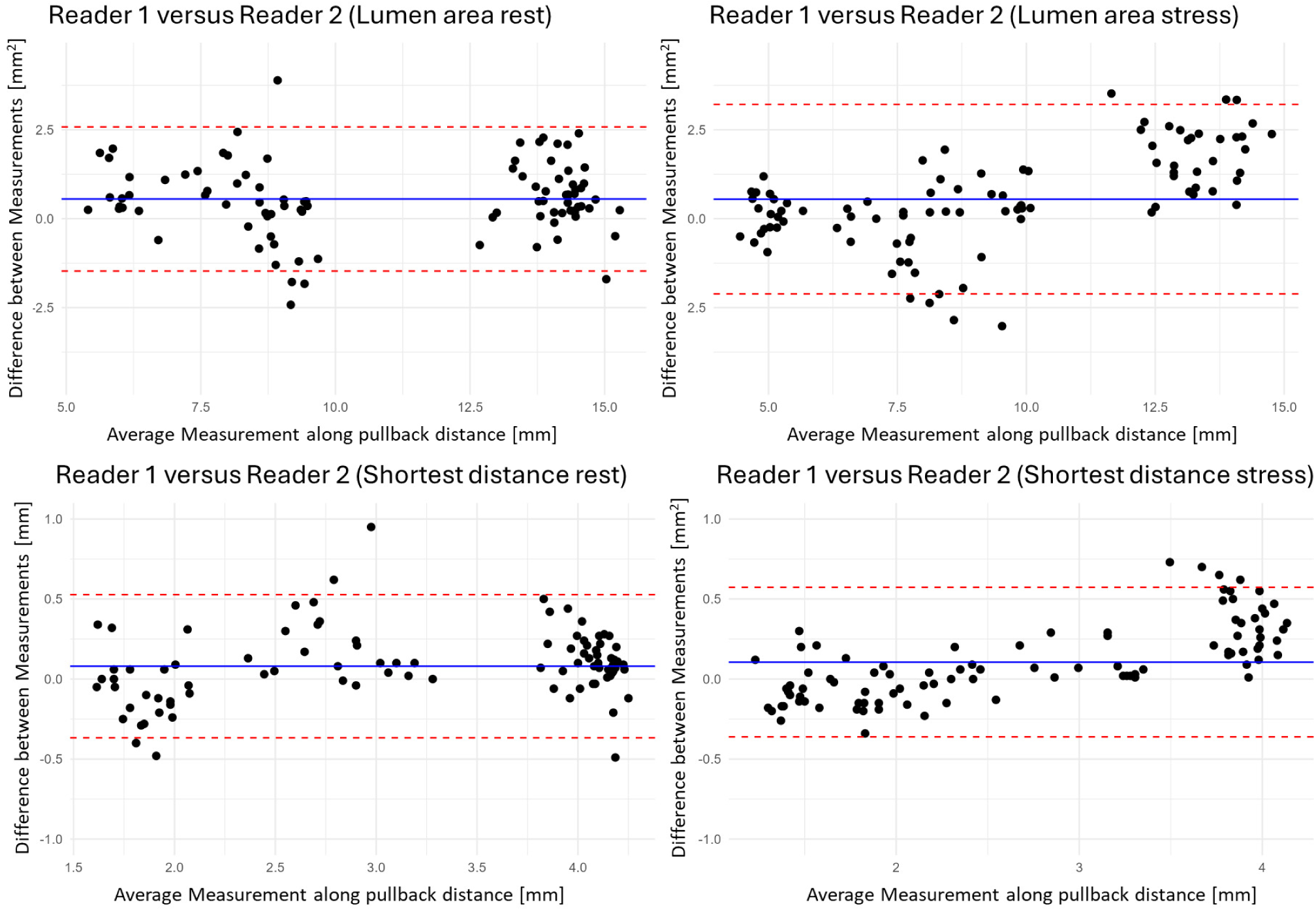
Bland-Altman plot for showing the difference between reader 1 and reader 2 for segmentation measurements. For area, the ICC was 0.94 (95%CI: 0.87-0.97, p < 0.001) for rest and 0.91 (95%CI: 0.84-0.94, p < 0.001) for stress. For shortest distance, the ICC was 0.97 (95%CI: 0.85-0.98, p < 0.001) for rest and 0.98 (95%CI: 0.97-0.99, p < 0.001) for stress.

**Figure A.12:**
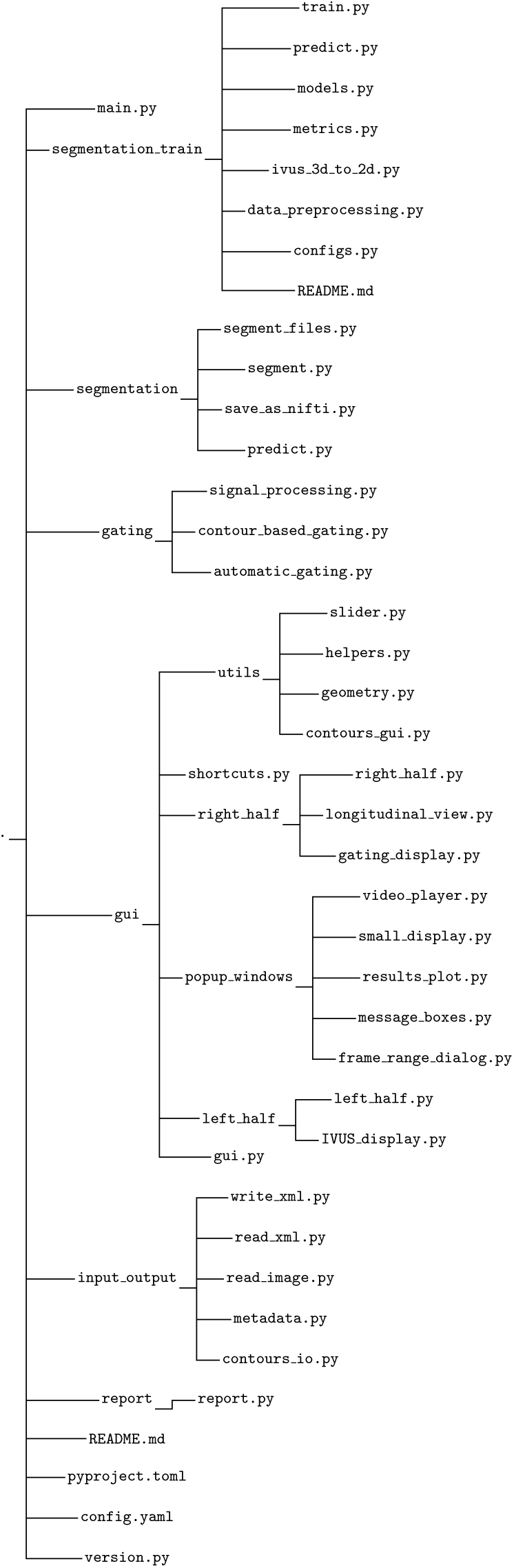
A visual representation of the project directory structure.

